# Repeated Hand Grip Strength is an Objective Marker for Disability and Severity of Key Symptoms in Post-COVID ME/CFS

**DOI:** 10.1101/2024.01.25.24301776

**Authors:** Anna Paffrath, Laura Kim, Claudia Kedor, Elisa Stein, Rebekka Rust, Helma Freitag, Uta Hoppmann, Leif G Hanitsch, Judith Bellmann-Strobl, Kirsten Wittke, Carmen Scheibenbogen, Franziska Sotzny

**Affiliations:** Institute of Medical Immunology, Charité – Universitätsmedizin Berlin, corporate member of Freie Universität Berlin and Humboldt-Universität zu Berlin, Augustenburger Platz 1, 13353 Berlin, Germany; Experimental and Research Center (ECRC), Charité – Universitätsmedizin Berlin, corporate member of Freie Universität Berlin and Humboldt-Universität zu Berlin, Charitéplatz 1, 10117 Berlin, Germany

**Keywords:** Myalgic Encephalomyelitis, Post Covid Syndrome, Chronic Fatigue Syndrome, Hand Grip Strength, Covid

## Abstract

Post-COVID Syndrome (PCS) refers to a diverse array of symptoms that persist beyond 3 months of the acute phase of a SARS-CoV-2 infection. The most frequent symptom is fatigue, which can manifest both mentally and physically. In this study, handgrip strength (HGS) parameters were determined as an objective measure of muscle fatigue and fatigability. HGS parameters were correlated with other frequent symptoms among 144 female PCS patients suffering from fatigue, exertional intolerance, and cognitive impairment. Seventy-eight patients met the Canadian Consensus Criteria (CCC) for post-infectious myalgic encephalomyelitis/chronic fatigue syndrome (ME/CFS). The severity of disability and key symptoms were evaluated utilizing self-reported questionnaires. Notably, patients diagnosed with ME/CFS exhibited a higher overall severity of symptoms, including lower physical function (*p* < 0.001), a greater degree of disability (*p* < 0.001), more severe fatigue (*p* < 0.001), post-exertional malaise (*p* < 0.001), and autonomic dysfunction (*p* = 0.004). While HGS was similarly impaired in both PCS and ME/CFS patients, the associations between HGS and the severity of symptoms and disability revealed striking differences. We observed significant correlations of HGS parameters with physical function across all patients, but with the key symptoms PEM, fatigue, cognitive impairment, and autonomic dysfunction in ME/CFS patients only. This points to a common mechanism for these symptoms in the ME/CFS subtype, distinct from that in other types of PCS. Further HGS provides an objective marker of disease severity in ME/CFS.

## 1. Introduction

Following coronavirus disease 2019 (COVID-19) a worrying number of patients experience a diverse range of symptoms persisting beyond the acute phase of the infection [1]. The World Health Organization (WHO) defined this post-COVID condition as symptoms occurring within three months following a SARS-CoV-2 infection, that persist for at least two months. These symptoms must affect everyday functioning and cannot be explained by an alternative diagnosis [2].

While some of these persistent symptoms may be caused by long-term inflammation and damages, there is little evidence for organ comorbidity in the majority of patients [3,4]. Typical symptoms include fatigue, exertional intolerance, post-exertional malaise (PEM), cognitive and autonomic dysfunction, myalgia, and headaches often referred to as post-COVID syndrome (PCS) [1]. In a subset of these patients, diagnostic criteria for post-infectious myalgic encephalomyelitis/chronic fatigue syndrome (ME/CFS) are met[5-8].

Post-infectious ME/CFS had affected around 3 million patients on the European continent alone, already in pre-pandemic times [9]. Given the high prevalence of COVID-19, the increase of ME/CFS is likely substantial. This poses probably the most significant problem of the pandemic for healthcare and society, as ME/CFS not only profoundly affects patients’ quality of life but also, in most cases, is a chronic disease with little knowledge and resources available for disease management [9].

In our observational study, patients diagnosed with post-COVID-ME/CFS generally show no improvement in their health throughout the second year of the disease, while patients with non-ME/CFS PCS display a more favourable prognosis [8]. Most PCS patients with fatigue who do not meet the diagnostic criteria for ME/CFS experience milder exertional intolerance with no or shorter PEM [5]. Pathomechanisms identified in both PCS and ME/CFS include inflammation, autoantibodies, endothelial dysfunction with hypoperfusion, and impaired mitochondrial function [4,10,11].

Diagnosis of ME/CFS is based on sets of diagnostic criteria, with the most frequently used being the Canadian Consensus Criteria (CCC) and the Institute of Open Medicine (IOM) criteria, which require the presence of fatigue, PEM, non-refreshing sleep, and cognitive impairment, and/or orthosthatic intolerance [12]. Handgrip strength (HGS) has been established as a reliable tool for assessing muscle fatigue in both patients with ME/CFS [13] and PCS [5].

Our study aimed to achieve a more comprehensive understanding of the symptom pattern of PCS and ME/CFS. Studying HGS as an objective tool to measure muscle fatigue, we aimed to establish correlations between HGS and functional impairment and symptom severity within the PCS and ME/CFS patient groups.

## 2. Materials and Methods

### 2.1. Patients

A total of 144 female patients with persistent fatigue, and other symptoms for at least 6 months following mild to moderate COVID-19 diagnosed with PCS according to the WHO criteria [2] were included in this study. Patients were diagnosed and recruited at the outpatient department for immunodeficiencies at the Institute of Medical Immunology at the Charité-Universitätsmedizin Berlin between April 2021 and December 2022. Diagnosis of ME/CFS was based on the Canadian Consensus Criteria (CCC) [14] and a minimum of 14 hours of PEM [15]. According to the CCC, PCS patients suffering from persistent fatigue after COVID-19 for at least 6 months were divided into a group with ME/CFS *(n* = 78) and non-ME/CFS (*n* = 66). Patients were excluded from this study in case of relevant comorbidities [9], preexisting fatigue, or evidence of organ dysfunction. All patients had to provide proof of previous SARSCoV-2 infection by positive PCR, antigen test, or serology (SARS-CoV-2 nucleocapsid protein antibodies).

All patients signed informed consent before study inclusion. This study was part of the Pa-COVID19 study of the Charité-Universitätsmedizin Berlin and was approved by the Ethics Committee of the Charité Universitätsmedizin Berlin in accordance with the 1964 Declaration of Helsinki and its later amendments (EA2/066/20 and EA2/067/20).

### 2.2. Hand Grip Strength Measurement

HGS of the dominant hand was measured using a digital hand dynamometer (Deyard ®, model: EH101) in two separate sessions. Rest time between sessions was 60 minutes, in which no strenuous physical activity took place. Patients sat in an upright position facing a standard table during measurements of HGS. The forearm of the dominant hand was placed on the table in full supination holding the dynamometer. Under supervision and verbal motivation, the handle was pulled 10 times with maximum force for three seconds, followed by a five-second relaxation phase. Before starting the measurement, the participants were shown two separate demonstrations of how the dynamometer should be used. The dynamometer displays the highest value reached within these three seconds (measurement in kg), and this single value is then recorded. The attempt with the highest reading out of ten repetitions was recorded as the maximum strength (Fmax) and the average force (Fmean) of each session was calculated. Fatigue Ratio (Fmax/Fmean) a measure of decrease of force during one session, and Recovery Ratio (Fmean2/Fmean1) a measure of recovery of force between both sessions were calculated [13].

### 2.3. Questionnaires for Symptom Scoring

Patients’ health-related quality of life was assessed using the 36-Item Short Form Survey (SF-36) ranging from 0 to 100 with 100 indicating no restriction [16]. Additionally, disease-related disability was scored according to the Bell score, rating the restriction in daily functioning on a scale from 0 to 100 with 100 indicating no restriction [17]. The severity of perceived fatigue was assessed using the Chalder Fatigue Scale (CFQ) ranging from 0 to 33 [18]. Post-exertional malaise (PEM) was evaluated using the DePaul Symptom Questionnaire (PEM-DSQ) ranging from 0 to 46 [15]. Further cardinal symptoms of both PCS and ME/CFS, including fatigue, muscle pain, immunological symptoms, and cognitive impairment, were scored on a scale from 1 to 10 [19,20]. The fatigue score was calculated as the mean of fatigue, malaise after exertion, need for rest, and daily functioning, and the cognitive score as a mean of memory disturbance, concentration ability, and mental tiredness based on symptoms recorded with the CCC. Autonomic dysfunction was evaluated according to the Composite Autonomic Symptom Score (COMPASS 31) ranging from 0 to 100 [21].

### 2.4. Statistical Analysis

Study data were collected and managed using REDCap electronic data capture tools hosted at Charité Universitätsmedizin Berlin [22,23].

Statistical data analyses were performed using IBM SPSS Statistics 29.0 (New York, NY, USA and R 4.3.1 (R Foundation for Statistical Computing, Vienna, Austria, http://www.R-project.org, accessed on 1 August 2023) and GraphPad Prism version 9 for Windows (GraphPad Software, San Diego, California USA, www.graphpad.com). All data are presented as the median and interquartile range, or frequency (*n*), and percentage where appropriate. Comparisons of quantitative parameters between two groups were performed using the nonparametric Mann–Whitney U test. Correlation analysis was performed using the nonparametric Spearman coefficient. The *p*-values < 0.05 were considered to provide evidence for a statistically significant result.

## Results

### 3.1. Patient characteristics

This analysis included a cohort of 144 female participants diagnosed with PCS. Among these patients, 78 fulfilled the diagnostic criteria of ME/CFS, while the remaining 66 participants did not. The demographic and clinical characteristics of the study population are presented in detail in Table 1.

**Table 1.** Demographic and clinical characteristics of the study population.

|  | **PCS-ME/CFS**<br>**(*n*=78, Median with range)** | **PCS-Non-ME/CFS**<br>**(*n*=66, Median with range)** | **ME/CFS vs.**<br>**non-ME/CFS** |
| --- | --- | --- | --- |
| Age | 40 (20-61) | 44 (18-67) | *p* = 0.585 |
| BMI | 23.71 (14.88-37.97) | 24.13 (17.65-41.64) | *p* = 0.675 |
| Time since COVID-19 (months) | 10 (6-25) | 10.5 (6-23) | *p* = 0.847 |
| Bell | 40 (20-90) | 50 (20-80) | *p* < 0.001 *** |
| CFQ | 28.5 (13-33) | 25 (0-33) | *p* < 0.001 *** |
| PEM-DSQ | 34 (12-46) | 28 (0-44) | *p* < 0.001 *** |
| SF-36 |  |  |  |
| Physical Functioning | 40 (0-95) | 60 (0-100) | *p* < 0.001 *** |
| Role Limitations | 0 (0-100) | 0 (0-100) | *p* = 0.107 |
| Energy | 15 (0-80) | 20 (0-60) | *p* < 0.001 *** |
| Pain | 32.5 (0-100) | 38.75 (0-100) | *p* = 0.064 |
| Symptom Severity |  |  |  |
| Muscle Pain | 6 (1-10) | 6 (1-10) | *p* = 0.324 |
| Headache | 7 (1-10) | 6 (1-10) | *p* = 0.115 |
| Joint Pain | 5 (1-10) | 6 (1-10) | *p* = 0.647 |
| Fatigue Score | 8 (1-10) | 7 (1-10) | *p* < 0.001 *** |
| Cognitive Score | 6.67 (1.33-10) | 6.5 (1-10) | *p* = 0.196 |
| Immune Score | 3.33 (1-8.67) | 2.33 (1-9) | *p* = 0.006 ** |
| COMPASS 31 | 40.80(0-76.23) | 30.64 (0-62.22) | *p* = 0.002** |
| Orthostatic Intolerance | 22 (0-40) | 16 (0-36) | *p* = 0.006 ** |
| Vasomotor | 0 (0-5) | 0 (0-3.33) | *p* = 0.003 ** |
| Secretomotor | 4.29 (0-12.86) | 4.29 (0-12.86) | *p* = 0.485 |
| Gastrointestinal | 6.25 (0-15.18) | 6.25 (0-17.86) | *p* = 0.187 |
| Bladder | 1.12 (0-7.78) | 1.11 (0-7.78) | *p* = 0.309 |
| Pupillomotor | 1.67 (0-4) | 1.67 (1-5) | *p* = 0.634 |
| Hand Grip Strength (in kg) |  |  |  |
| Fmax 1 | 20.1 (5.7-43.6) | 19.2 (4.7-33.8) | *p* = 0.884 |
| Fmax 2 | 18.25 (1.9-41.2) | 17.1 (3.10-34.6) | *p* = 0.477 |
| Fmean 1 | 16.6 (3.19-38.99) | 16.07 (4.14-31.85) | *p* = 0.938 |
| Fmean 2 | 15.07 (1.37-39.05) | 14.51 (2.32-30.88) | *p* = 0.990 |
| Fatigue Ratio 1 | 1.18 (1.01-3.23) | 1.17 (1.03-2.11) | *p* = 0.642 |
| Fatigue Ratio 2 | 1.2 (1.04-2.24) | 1.17 (1.03-2.0) | *p* = 0.035 * |
| Recovery Ratio | 0.95 (0.26-1.67) | 0.95 (0.47-1.24) | *p* = 0.674 |

Asterisks mark significant differences between groups (Mann-Whitney test, \* *p* < 0.05, \*\* *p* < 0.01, \*\*\* *p* < 0.001 \*\*\*). BMI, Body Mass Index; CFQ; Chalder Fatigue Scale; COMPASS 31; Composite Autonomic Symptom Score; Fatigue Ratio 1/2, Ratio of Fmax/Fmean of the first/second session; Fmax 1/2, maximum hand grip strength of the first/second session; Fmean 1/2, mean hand grip strength of the first/second session; ME/CFS, Myalgic Encephalomyelitis/Chronic Fatigue Syndrome; PCS, Post-COVID Syndrome; PEM-DSQ; De Paul Symptom Questionnaire for Post Exertional Malaise; Recovery Ratio, Ratio of Fmean2/Fmean1, SF-36, Short-Form 36 Health Survey

### 3.1. Comparison between Patient Groups

As shown in Table 1, PCS patients across both patient groups displayed a broad spectrum of symptoms, not limited to fatigue, and PEM: headaches, arthralgia, myalgia, cognitive dysfunction, and autonomic dysfunction, and others.

As depicted in Table 1 and Figure 1 patients diagnosed with ME/CFS exhibited significantly lower physical function in the SF-36 (*p* < 0.001) and had a significantly higher degree of disability attributed to their illness as assessed via the Bell score (*p* < 0.001) in comparison to PCS without ME/CFS. Moreover, individuals with ME/CFS exhibited more severe orthostatic intolerance (*p* = 0.006), vasomotor symptoms (*p*= 0.003) fatigue (*p* < 0.001), and PEM (*p* < 0.001).

**Figure 1.**
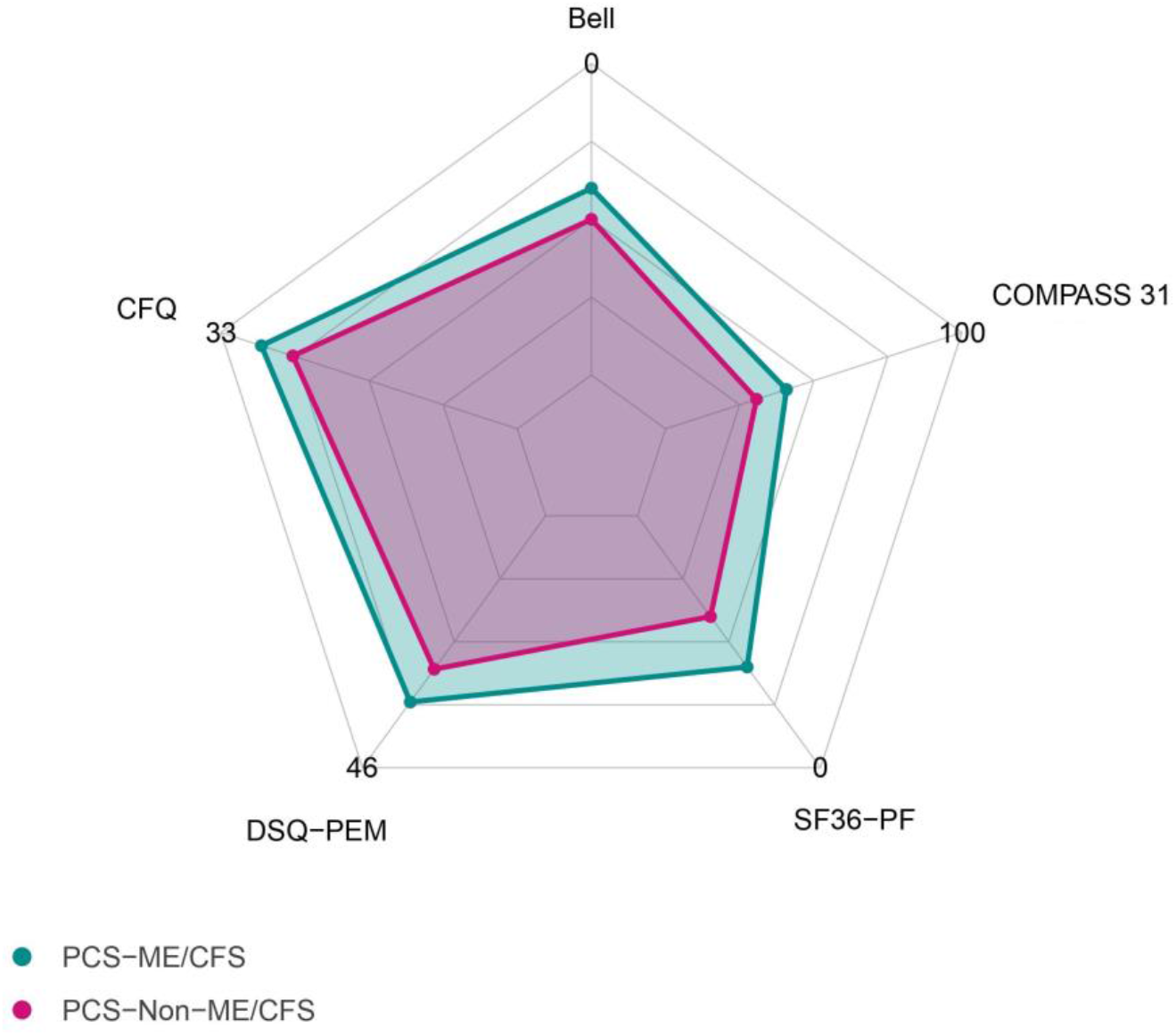
Median scores of questionnaires in patients diagnosed with either PCS or ME/CFS. Bell, disease-related disability according to the Bell-Score; CFQ, Chalder Fatigue Questionnaire; COMPASS 31, Composite Autonomic Symptom Score; DSQ-PEM, De Paul Symptom Questionnaire for Post Exertional Malaise; SF-36-PF, Short-Form 36 Health Survey Physical Function

### 3.2. Hand Grip Strength

Adjusted for age and gender, the median maximum HGS (Fmax 1) in PCS patients across both groups was below the normal values for the general population provided by the dynamometer manufacturer. Further, we calculated the mean HGS for the initial set of 10 measurements (Fmean 1) and the set taken after a one-hour interval (Fmean 2) and Fatigue Ratios (Fmax/Fmean) as a parameter of fatigability within the set of 10 measurements as described by Jäkel *et al* [13]. The Recovery Ratio was calculated as (Fmean 2/ Fmean 1) with values below 1 indicating impaired recovery after the one-hour interval.

As shown in Table 1, there were little differences in HGS parameters observed between the two patient cohorts with only Fatigue Ratio 2 being lower in ME/CFS patients.

As illustrated in Figure 2 and Table S1, the parameters of HGS demonstrated significant correlations with various parameters of functional disability and symptom severity among patients diagnosed with ME/CFS.

**Figure 2.**
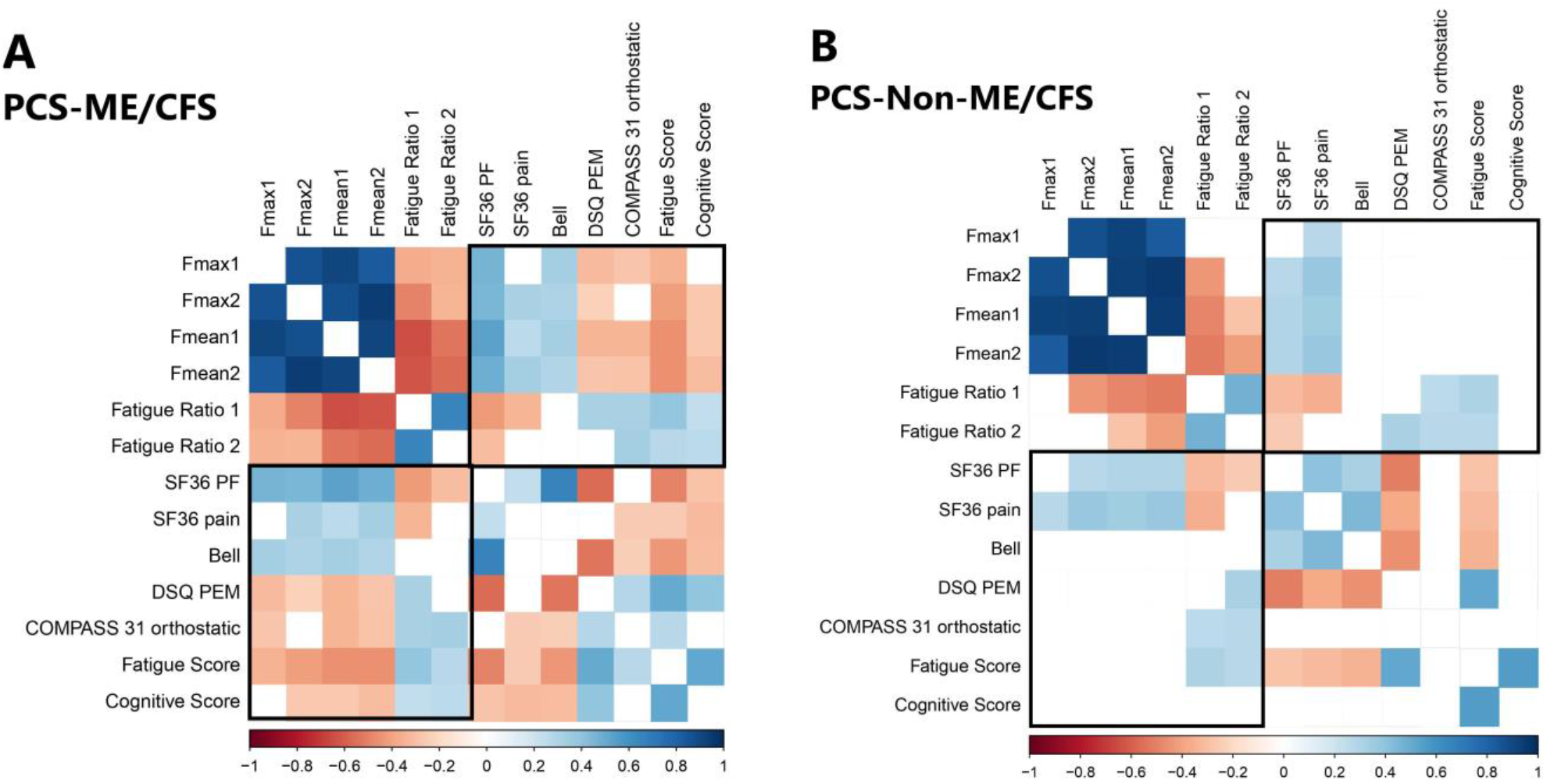
Correlation matrix showing correlation coefficients of parameters of symptom severity and handgrip strength (HGS) for both patient groups. Rectangles indicate the parameters of HGS. Nonsignificant coefficients were left blank. (**A**) Patients diagnosed with both PCS and ME/CFS; (**B**) patients diagnosed with PCS but not ME/CFS.

Notably, there were strong associations between HGS parameters and PEM as well as measures of disability, as evaluated by the Bell score, and the SF-36 physical function in ME/CFS patients. Additionally, correlations were observed between HGS parameters and fatigue, cognitive impairment, pain, and orthostatic intolerance assessed by the COMPASS score (Figure 2). It is important to note that in PCS patients without ME/CFS a similar correlation of the HGS parameters was only found with SF-36 physical function and pain. Furthermore, while fatigue, pain, and PEM correlated with functional disability in both patient cohorts, correlations of cognition and orthostatic intolerance with functional disability and PEM were observed only in ME/CFS but not non-ME/CFS patients, despite similar severity.

## 4. Discussion

Our objective was to elucidate potential correlations between HGS parameters, disability, and key symptoms. HGS was previously shown to be a sensitive diagnostic test to assess muscular fatigue and fatigability in patients with ME/CFS [13]. Further, lower HGS was associated with a lower Bell score, more PEM, and myalgia in prepandemic postinfectious ME/CFS [13]. In PCS lower HGS 6 months after disease onset was a risk factor for chronification [8].

In this study, we observed a diverse range of symptoms among our PCS cohort in line with previous studies [5,8]. Patients with ME/CFS had more severe fatigue, PEM, and orthostatic intolerance. Furthermore, ME/CFS patients experienced substantially greater disability due to their illness and demonstrated lower levels of physical functioning overall. This observation underscores that, although the range of symptom manifestation might be similar between these conditions, Post-COVID-ME/CFS is a more severe form of PCS. The diagnosis of ME/CFS following a COVID-19 infection is not only linked to an increased symptom burden and elevated risk of disability but it also brings about a heightened risk of chronification, as shown in our prior study [8].

We found that HGS was impaired in all PCS patients, irrespective of ME/CFS diagnosis. However, while SF36 physical function and pain correlated with diminished HGS in all patients, correlations with PEM, cognition, fatigue score, and orthostatic intolerance were only found within the ME/CFS patient group. Most striking is the absence of a correlation between HGS and cognitive impairment in the non-ME/CFS PCS subtype, despite similar symptom severity. Further, cognition correlated with functional disability, pain, and PEM only in ME/CFS but not non-ME/CFS patients.

This points to a common mechanism of muscle fatigue, general fatigue, cognitive impairment, and orthostatic intolerance in ME/CFS which is distinct from non-ME/CFS PCS. Several studies have elucidated mechanisms of muscle fatigue, orthostatic intolerance, and cognitive impairment. A central finding in both ME/CFS and PCS is cardiovascular dysfunction and hypoperfusion as shown by impaired cerebral blood flow and systemic oxygen extraction [24]. The cause for impaired perfusion in ME/CFS is complex and can be related to both “impaired cardiac preload” and “microcirculatory dysfunction”. Potential mechanisms include dysautonomia, endothelial dysfunction, dysfunctional red blood cells, and possibly microclots. Inflammation and mitochondrial impairment can aggravate muscle fatigue. In line with this, histological studies of muscles in PCS revealed capillary injury and rarefication, mitochondrial changes, and inflammation [25,26]. A MRI study revealed higher sodium content in muscles of ME/CFS patients compared to healthy controls and a correlation with HGS suggesting impaired function of ion transport presumably due to hypoperfusion [27].

Our findings suggest that despite the presence of common symptoms across the group of patients with PCS, ME/CFS may have distinct or additional mechanisms. We have first evidence for this notion from biomarker studies. We found that HGS was associated with distinct biomarker profiles suggesting an association with inflammation in PCS and perfusion and muscle damage in ME/CFS [5]. Previously, we found an association between fatigue and impaired peripheral perfusion with autoantibodies to vasoregulatory receptors in ME/CFS but not PCS [10]. We also observed differences in endothelial biomarkers and a proangiogenic pattern in PCS but not ME/CFS [28]. These findings provide first evidence that mechanisms of muscle fatigue and vascular dysfunction are probably distinct in ME/CFS.

Importantly our study provides evidence that repeated HGS is an objective marker of physical function in PCS patients, and also of severity of key symptoms in the ME/CFS subgroup. Thus, repeated HGS is an important diagnostic and prognostic tool allowing an objective assessment of disability.

Notably, our study is also constrained by limitations, including that the PCS patients constitute a not well-defined subgroup characterized by moderate to severe fatigue, exertion intolerance, and cognitive impairment and the absence of control groups, particularly of recovered post-COVID-19 individuals. Moreover, this study exclusively involved female patients.

## 5. Conclusions

Patients with PCS can present with a broad spectrum of symptoms. HGS impairment was evident in PCS patients with no difference in the ME/CFS subgroup. HGS is an important measure of muscle fatigue and correlates with disability and symptom severity in ME/CFS thus providing an objective parameter of disease severity. Associations between symptoms and HGS impairment found predominantly in the ME/CFS subgroup may indicate a distinct mechanism or mechanistic link. Thus, the subgrouping of PCS patients according to ME/CFS criteria in biomarker studies is important due to potential differences in underlying pathomechanisms.

## Supplementary Materials

Table S1: Correlations of HGS parameters and clinical parameters.

## Author Contributions

Conceptualization C.S. and F.S.; methodology, F.S. and H.F.; validation, F.S and L.K.; formal analysis, A.P. and L.K.; investigation, A.P., C.K., E.S., R.R., U.H., L.G.H. J.B.-S., K.W. and C.S.; resources, C.S.; data curation H.F., F.S. and A.P.; writing—original draft preparation, L.K.; writing—review and editing, C.S. and F.S.; visualization, L.K.; supervision, C.S. and F.S.; project administration, F.S.. All authors have read and agreed to the published version of the manuscript.

## Institutional Review Board Statement

The study was conducted in accordance with the Declaration of Helsinki, and approved by the Ethics Committee of the Charité - Universitätsmedizin Berlin (EA2/066/20 from 08.04.2021 and EA2/067/20 from 27.04.2020).

## Informed Consent Statement

Informed consent was obtained from all subjects involved in the study.

## Data Availability Statement

The data presented in this study are available on request from the corresponding author.

## Acknowledgments

We thank Anja Hagemann, Silvia Thiel, and Beate Follendorf for patient care and data management. We thank all patients who gave us their consent. We thank all members of the Pa-COVID-19 collaborative study group.

