## Supplemental Table 1 for "Repeated Hand Grip Strength is an Objective Marker for Disability and Severity of Key Symptoms in Post-COVID ME/CFS"

**Table S1.** Correlations of HGS parameters and clinical parameters.

| | Group | Fmax 1 | Fmax 2 | Fmean 1 | Fmean 2 | Fatigue Ratio 1 | Fatigue Ratio 2 | Recovery Ratio |
|---|---|---|---|---|---|---|---|---|
| Bell | PCS-ME/CFS | *r* = 0.34** | *r* = 0.31** | *r* = 0.33** | *r* = 0.29* | *r* = -0.2 | *r* = -0.14 | *r* = 0.14 |
| | PCS-Non-ME/CFS | *r* = 0.19 | *r* = 0.17 | *r* = 0.17 | *r* = 0.12 | *r* = -0.13 | *r* = 0.01 | *r* = -0.08 |
| CFQ | PCS-ME/CFS | *r* = -0.23* | *r* = -0.19 | *r* = -0.26* | *r* = -0.2 | *r* = 0.16 | *r* = 0.14 | *r* = 0.04 |
| | PCS-Non-ME/CFS | *r* = -0.2 | *r* = -0.18 | *r* = -0.21 | *r* = -0.2 | *r* = 0.23 | *r* = 0.19 | *r* = -0.08 |
| DSQ-PEM | PCS-ME/CFS | *r* = -0.32** | *r* = -0.24* | *r* = -0.33** | *r* = -0.27* | *r* = 0.32** | *r* = 0.23 | *r* = -0.02 |
| | PCS-Non-ME/CFS | *r* = -0.23 | *r* = -0.23 | *r* = -0.27 | *r* = -0.27 | *r* = 0.24 | *r* = 0.32* | *r* = -0.07 |
| SF-36 | | | | | | | | |
| Physical Function | PCS-ME/CFS | *r* = 0.47*** | *r* = 0.45*** | *r* = 0.53*** | *r* = 0.48*** | *r* = -0.43*** | *r* = -0.31** | *r* = 0.16 |
| | PCS-Non-ME/CFS | *r* = 0.24 | *r* = 0.27* | *r* = 0.3* | *r* = 0.3* | *r* = -0.31* | *r* = -0.26* | *r* = 0.08 |
| Role Limitations | PCS-ME/CFS | *r* = 0.16 | *r* = 0.2 | *r* = 0.21 | *r* = 0.22 | *r* = -0.26* | *r* = -0.20 | *r* = 0.12 |
| | PCS-Non-ME/CFS | *r* = 0.29* | *r* = 0.3* | *r* = 0.35** | *r* = 0.33** | *r* = -0.3* | *r* = -0.29* | *r* = 0.07 |
| Energy | PCS-ME/CFS | *r* = 0.3** | *r* = 0.38*** | *r* = 0.34** | *r* = 0.39*** | *r* = -0.32** | *r* = -0.13 | *r* = 0.13 |
| | PCS-Non-ME/CFS | *r* = 0.14 | *r* = 0.12 | *r* = 0.19 | *r* = 0.15 | *r* = -0.26* | *r* = -0.24* | *r* = -0.007 |
| Pain | PCS-ME/CFS | *r* = 0.19 | *r* = 0.32** | *r* = 0.26* | *r* = 0.33** | *r* = -0.33** | *r* = -0.22 | *r* = 0.24* |
| | PCS-Non-ME/CFS | *r* = 0.28* | *r* = 0.38** | *r* = 0.34** | *r* = 0.37** | *r* = -0.35** | *r* = -0.17 | *r* = 0.1 |
| COMPASS 31 | PCS-ME/CFS | *r* = -0.21 | *r* = -0.15 | *r* = -0.26* | *r* = -0.22 | *r*= 0.3* | *r* = 0.3* | *r* = -0.03 |
| | PCS-Non-ME/CFS | *r* = -0.02 | *r* = 0.05 | *r* = -0.03 | *r* = -0.04 | *r* = 0.32** | *r* = 0.22* | *r* = -0.13 |
| Orthostatic | PCS-ME/CFS | *r* = -0.27* | *r* = -0.2 | *r* = -0.34** | *r* = -0.29* | *r* = 0.33** | *r* = 0.34** | *r* = -0.11 |
| | PCS-Non-ME/CFS | *r* = -0.01 | *r* = 0.03 | *r* = -0.06 | *r* = -0.07 | *r* = 0.27* | *r* = 0.27* | *r* = -0.14 |
| Vasomotor | PCS-ME/CFS | *r* = 0.03 | *r* = 0.02 | *r* = -0.06 | *r* = -0.03 | *r* = 0.15 | *r* = 0.15 | *r* = 0.04 |
| | PCS-Non-ME/CFS | *r* = 0.03 | *r* = 0.03 | *r* = 0.04 | *r* = 0.01 | *r* = -0.01 | *r* = 0.01 | *r* = -0.13 |
| Secretomotor | PCS-ME/CFS | *r* = 0.29* | *r* = 0.28* | *r* = 0.21 | *r* = 0.25* | *r* = -0.12 | *r* = -0.13 | *r* = 0.13 |
| | PCS-Non-ME/CFS | *r* = -0.15 | *r* = -0.1 | *r* = -0.14 | *r* = -0.1 | *r* = 0.04 | *r* = 0.05 | *r* = 0.11 |
| Gastrointestinal | PCS-ME/CFS | *r* = 0.19 | *r* = 0.16 | *r* = 0.16 | *r* = 0.19 | *r* = -0.12 | *r* = -0.1 | *r* = 0.06 |
| | PCS-Non-ME/CFS | *r* = 0.15 | *r* = 0.15 | *r* = 0.14 | *r* = 0.16 | *r* = 0.03 | *r* = -0.03 | *r* = 0.22 |
| Bladder | PCS-ME/CFS | *r* = -0.15 | *r* = -0.09 | *r* = -0.1 | *r* = -0.1 | *r* = -0.08 | *r* = 0.13 | *r* = -0.04 |
| | PCS-Non-ME/CFS | *r* = -0.04 | *r* = 0.01 | *r* = -0.04 | *r* = -0.02 | *r* = 0.1 | *r* = 0.17 | *r* = 0.16 |
| Pupillomotor | PCS-ME/CFS | *r* = 0.26* | *r* = 0.35** | *r* = 0.27* | *r* = 0.32** | *r* = -0.28* | *r* = 0.9 | *r* = 0.16 |
| | PCS-Non-ME/CFS | *r* = 0.04 | *r* = 0.01 | *r* = 0.08 | *r* = 0.04 | *r* = -0.08 | *r* = -0.02 | *r* = -0.08 |
| Symptom Severity | | | | | | | | |
| Muscle pain | PCS-ME/CFS | *r* = -0.2 | *r* = -0.22 | *r* = -0.26* | *r* = -0.27* | *r* = 0.24* | *r* = 0.3 ** | *r* = -0.05 |
| | PCS-Non-ME/CFS | *r* = -0.1 | *r* = -0.16 | *r* = -0.17 | *r* = -0.19 | *r* = 0.21 | *r* = 0.23 | *r* = -0.02 |
| Headache | PCS-ME/CFS | *r* = -0.06 | *r* = -0.12 | *r* = -0.06 | *r* = -0.1 | *r* = 0.05 | *r* = 0.09 | *r* = -0.02 |
| | PCS-Non-ME/CFS | *r* = -0.04 | *r* = -0.1 | *r* = -0.11 | *r* = -0.06 | *r* = 0.32** | *r* = -0.07 | *r* = 0.1 |
| Joint pain | PCS-ME/CFS | *r* = -0.33** | *r* = -0.34** | *r* = -0.43*** | *r* = -0.41*** | *r* = 0.43*** | *r* = 0.36** | *r* = -0.09 |
| | PCS-Non-ME/CFS | *r* = -0.009 | *r* = -0.17 | *r* = -0.13 | *r* = -0.19 | *r* = 0.32* | *r* = 0.24 | *r* = -0.07 |
| Fatigue Score | PCS-ME/CFS | *r* = -0.35** | *r* = -0.42** | *r* = -0.46** | *r* = -0.46** | *r* = 0.39** | *r* = 0.28* | *r* = -0.07 |
| | PCS-Non-ME/CFS | *r* = -0.044 | *r* = -0.06 | *r* = -0.11 | *r* = -0.11 | *r* = 0.32* | *r* = 0.27* | *r* = -0.02 |
| Cognitive Score | PCS-ME/CFS | *r* = -0.22 | *r* = -0.26* | *r* = -0.27* | *r* = -0.3** | *r* = 0.24* | *r* = 0.28* | *r* = -0.17 |
| | PCS-Non-ME/CFS | *r* = -0.2 | *r* = -0.15 | *r* = -0.16 | *r* = -0.12 | *r* = 0.09 | *r* = 0.05 | *r* = 0.03 |
| Immune Score | PCS-ME/CFS | *r* = -0.16 | *r* = -0.26* | *r* = -0.24* | *r* = -0.31** | *r* = 0.24* | *r* = 0.34** | *r* = -0.17 |
| | PCS-Non-ME/CFS | *r* = -0.008 | *r* = -0.04 | *r*=- 0.03 | *r* = -0.09 | *r* = 0.3* | *r* = 0.25 | *r* = -0.14 |

Asterisks mark significant correlations (Spearman Correlation, * p < 0.05, ** p < 0.01, ***p < 0.001 ***).

CFQ; Chalder Fatigue Scale; COMPASS 31; Composite Autonomic Symptom Score; Fatigue Ratio 1/2, Ratio of Fmax/Fmean of the first/second session; Fmax 1/2, maximum hand grip strength of the first/second session; Fmean 1/2, mean hand grip strength of the first/second session; ME/CFS, Myalgic Encephalomyelitis/Chronic Fatigue Syndrome; PCS, Post-COVID Syndrome; PEM-DSQ; De Paul Symptom Questionnaire for Post Exertional Malaise; Recovery Ratio, Ratio of Fmean2/Fmean1, SF-36, Short-Form 36 Health Survey
